# The Fontan Dapagliflozin Pilot Study (FonDap)

**DOI:** 10.64898/2026.06.23.26356392

**Authors:** Ari Cedars, Gentian Lluri, Jong Mi Ko, Avinash Dhimal, Rawan Amir, Lisa Yanek, Stacy Fisher, Jamil Aboulhosn

## Abstract

**Background:** Patients with Fontan are prone to sequelae related to chronic elevations in central venous pressures. Interventions that improve venous pressures without compromising ventricular filling may therefore be of benefit.

**Methods:** We conducted a multi-center, open label, single arm pilot study of 4 weeks of dapagliflozin 10mg in adult patients with Fontan. The primary outcome was change in resting peripheral venous pressure (PVP). Secondary outcomes included changes in post-exercise PVP, peak VO2, Ve/VCO2, oxygen pulse, oxygen uptake efficiency slope (OUES), total body water, and patient reported health status according to the ACHD PRO.

**Results:** The total of 29 patients were enrolled between 11/1/2023 and 2/3/2026 across 2 centers, of whom 26 completed all study procedures. Average age was 31.2 years and 19 had a morphologic left ventricle. Dapagliflozin decreased PVP by 1.3mmHg (IQR −2.6, 0.4, p=0.012) with a greater effect in those with higher baseline PVP. Dapagliflozin improved patient reported health status and resulted in a trend towards an improvement in peak VO2 (0.2ml/kg/min, IQR −0.9, 2.2, p=0.064) and oxygen pulse (0.3ml/beat, IQR −0.6-1.1, p=0.074) without any impact on other cardiopulmonary exercise test parameters or total body water. Dapagliflozin was well tolerated in participants with no significant adverse events.

**Conclusions:** Dapagliflozin decreased PVP and improved patient reported health status in adult patients with Fontan over 4 weeks of therapy and was generally well tolerated.

## Introduction

Patients with Fontan are prone to multiple health problems related to their palliation. The great majority of these sequelae result directly or indirectly from chronically elevated central venous pressures which ultimately compromise longevity^1^. Physiologically, however, the Fontan circulation necessitates elevated venous pressures to overcome the resistance of the pulmonary vascular bed and maintain ventricular filling^2^. As patients with Fontan age, the onset of ventricular systolic or diastolic dysfunction leads to further elevation in venous pressures accelerating physiologic deterioration and leading ultimately to Fontan circulatory failure.

Sodium-glucose cotransporter-2 inhibitors (SGLT2i) are a mainstay of pharmacologic therapy for both systolic and diastolic heart failure in patients with normal cardiac anatomy^3^. Although the mechanisms underlying the benefits of SGLT2i are the subject of ongoing research, these drugs are known to decrease total body water and improve diuretic responsiveness^4,5^. Given the demonstrated benefits of SGLT2i in myocardial disease with fluid overload, these drugs are of interest as potentially beneficial pharmacologic interventions for patients with Fontan. Recent case reports and series hint at potential benefit^6–8^, however SGLT2i have yet to be systematically studied in the Fontan population. To begin filling this knowledge gap, we conducted an open-label pilot study to investigate the subacute physiologic effects of the SGLT2i dapagliflozin in patients with Fontan.

## Methods

We conducted a multicenter open-label pilot study of dapagliflozin in patients with Fontan. The study protocol and conduct were approved by the institutional review boards at Johns Hopkins School of Medicine and the University of California in Los Angeles, the two sites at which the study was conducted. This study was funded by the Bret Boyer Foundation and Astra Zeneca Pharmaceutical Company provided the study drug dapagliflozin. The study was registered with ClinicalTrials.gov with a reference number NCT05741658.

### Study Population

We aimed to enroll 29 patients actively followed at participating institutions aged 18 years or older who were physically able to complete an ergometer-based cardiopulmonary exercise test and undergo bioimpedance testing for total body water composition assessment; and who were able to read, understand and consent to the study protocol and complete the ACHD PRO V1 patient-reported outcome metric. Any patient who was unstable (defined as having had a procedure or being hospitalized in the previous 6 months); pregnant, planning to become pregnant or actively breast feeding; had obstruction in their Fontan circulation; was already treated with or had a prior adverse reaction to dapagliflozin; or had Type 1 diabetes, hypotension (systemic systolic blood pressure of < 95 mmHg) or significant impairment in their liver (alanine or aspartate amino transferase of >3x the upper limit of normal) or kidney function (Estimated glomerular filtration rate of <25ml per minute per 1.73m2 body surface area as assessed by MDRD calculation) were excluded..

### Study Outcomes

The primary outcome was change in pre-exercise peripheral venous pressure (PVP) as measured by peripheral venous manometry between study beginning and study end. Secondary outcomes included changes in post-exercise PVP, peak VO2, Ve/VCO2, oxygen pulse, and oxygen uptake efficiency slope (OUES) assessed during cardiopulmonary exercise testing; total body water composition as assessed using bioimpedance testing; and ACHD PRO V1 score between baseline and study end.

Laboratory values including complete blood count, comprehensive metabolic panel, cystatin C and NT-pro BNP were also monitored throughout the study period although they were not included as outcomes.

### Study Protocol

Sequential patients seen at the two participating sites who had an interest in participating in the study and met eligibility criteria were enrolled in the study between November 2023 and March 2026. Patients had a baseline visit at which time all outcome metrics were assessed. All patients then received dapagliflozin 10mg which they took once daily for 4 weeks. At the end of the 4-week period, patients returned for a repeat assessment of all outcome metrics. A complete study protocol including a study event table can be found in Supplemental Materials.

### Safety and Compliance Monitoring

An in-person safety monitoring visit was conducted at baseline, one week after starting dapagliflozin, at the 4-week study visit and one week after stopping dapagliflozin. In addition, a safety monitoring call was conducted the second and third weeks after starting dapagliflozin. Participants were each provided with an automatic cuff-style, upper-arm blood pressure and pulse monitor, verified for accuracy at the first study visit, to use at home to check twice daily blood pressures and record them for review at the time of study visits. Pills were counted at each in-person visit to ensure compliance. Adverse events were defined as any effect of the study drug or protocol which is unfavorable, undesirable, uncomfortable, or caused a participant distress and which deviated from the participant’s normal baseline symptom burden in the opinion of the participant. Serious adverse events were defined as any participant hospitalization, emergency room visit or death that took place during the study period as well as any change in participant clinical status or laboratory value which, in the opinion of the site PI, required immediate cessation of study drug.

### Statistical Analysis

Descriptive statistics including frequencies and proportions, means and standard deviations, or medians with 25th and 75th percentiles were calculated for the cohort’s characteristics of interest at baseline and 4-week follow-up. Change from baseline to follow-up was calculated as the follow-up value minus the baseline value. The Shapiro-Wilks test was used to test distributions of continuous variables for normality; normally distributed variables are presented as means with standard deviations, non-normally distributed variables are presented as medians with 25th and 75th percentiles. Within-patient changes are presented as means with 95% confidence limits and were tested with paired t-tests for normally distributed variables or signed-rank tests for non-normally distributed variables. The primary outcome was examined in stratified analysis by subgroups of interest. The primary and secondary outcomes were examined by baseline PVP >= median or < median and visualized in forest plots. All analyses were performed using SAS v 9.4 (SAS Institute, Cary, NC). P-values < 0.05 were considered to indicate statistical significance.

## Results

### Population

Target enrollment based on initial power analysis was 29 subjects allowing for 10% dropout. Twenty-nine subjects consented to participate in the study, 16 from Johns Hopkins University and 13 from the University of California Los Angeles. One subject was withdrawn as a liver lesion was identified the week prior to the first visit. Twenty-eight subjects started taking the study medication. Two subjects withdrew from the trial after medication administration due to possible medication side effects (one due to erectile dysfunction and one due to genital discharge, rash and fatigue). Twenty-six subjects completed the study protocol. Table 1 has descriptive statistics on the 28 patients who received study medication. There were no medication changes during the study protocol in any patient. Other medications taken during the study period are in Supplemental Table 1.

**Table 1:**
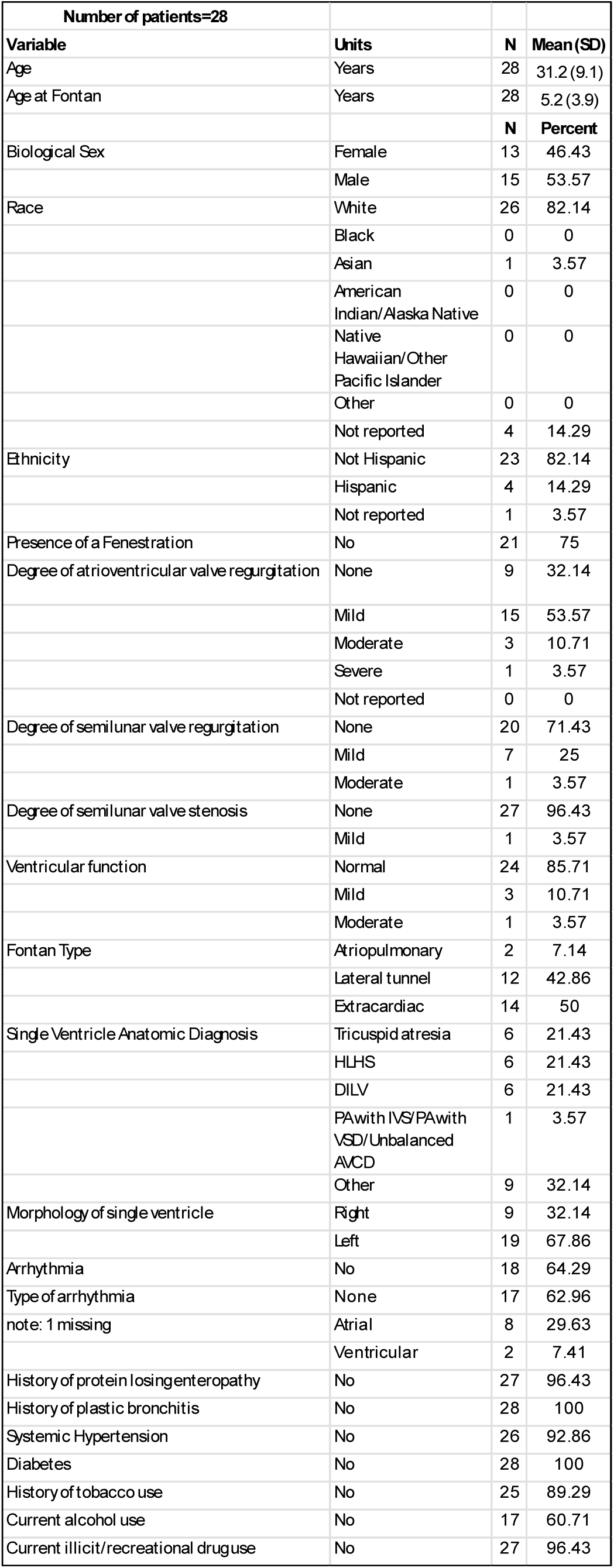
HLHS: Hypoplastic left heart syndrome; DILV: Double inlet left ventricle; PA: Pulmonary atresia; IVS/PA: Pulmonary atresia with intact ventricular septum; VSD: Ventricular septal defect; AVCD: Atrioventricular canal defect; PDE5: Type 5 phosphodiesterase; ACE: Angiotensin converting enzyme; ARB: Angiotensin receptor blocker

### Primary Outcome

Median baseline pre-exercise PVP was 9.2mmHg (IQR 7.4, 11.8) and decreased to 7.9mmHg (IQR 6.6, 10.9; p=0.01; Table 2). Although the decrease was seen across most groups, there was little effect of dapagliflozin in those with a PVP of <10mmHg and in those with abnormal ventricular function (Figure 1). When divided according to median baseline PVP, we found a decrease in PVP (−1.9mmHg [−3.2mmHg - −0.7mmHg]) in those with a PVP ≥ the median (9.6mmHg) while we found no change in those with PVP < median (−0.3mmHg [−1.5mmHg – 0.9mmHg] Figure 2).

**Figure 1:**
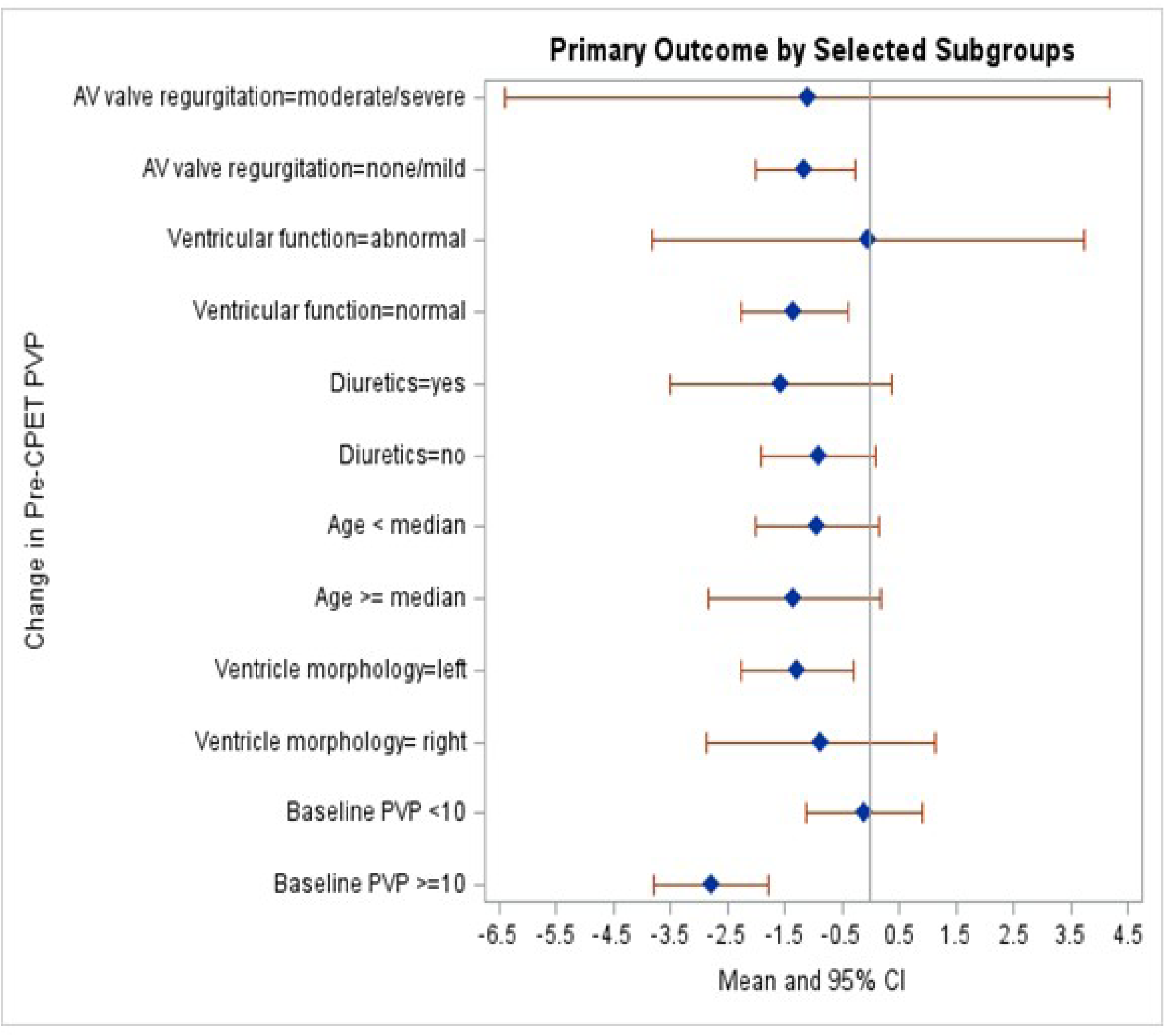
Forest plot depicting the change in peripheral venous pressure (PVP) in selected subgroups. AV: atrioventricular.

**Figure 2:**
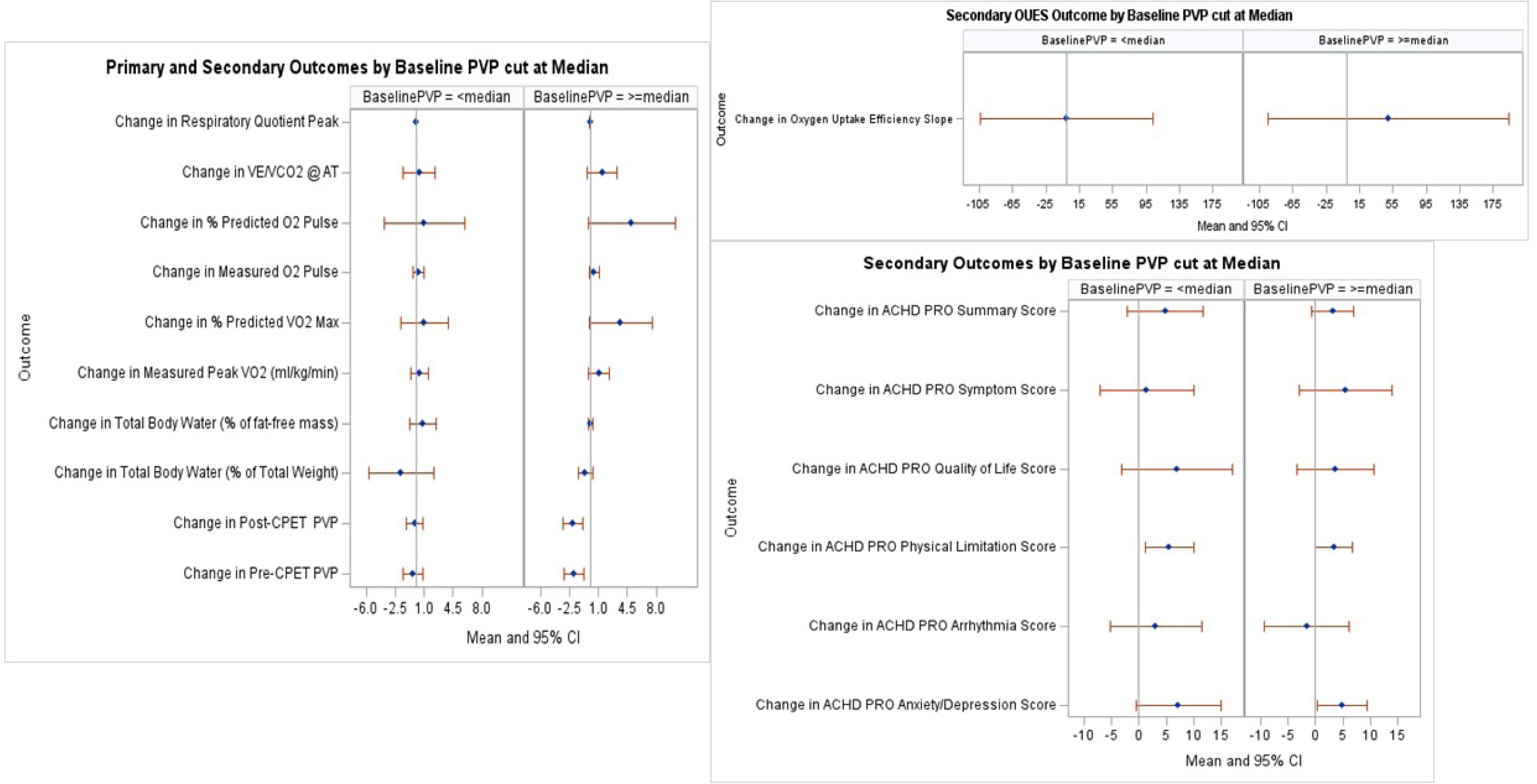
Forest plots depicting differences in all outcomes according to baseline peripheral venous pressure (PVP) at above or below median. Ve/VCO2: Ventilatory efficiency slope; AT: Anaerobic threshold; VO2: Oxygen uptake; CPET: Cardiopulmonary exercise test; OUES: Oxygen uptake efficiency slope.

**Table 2:** Primary and Secondary Outcomes; Significant values are indicated by italics.

| Primary and Secondary Outcomes:<br>Table 2 |  | Baseline |  |  | Follow-up |  |  | Change |  |  |  |  |  |  |
| --- | --- | --- | --- | --- | --- | --- | --- | --- | --- | --- | --- | --- | --- | --- |
|  | units | N | Mean (SD) | Median (25th, 75th %iles) | N | Mean (SD) | Median (25th, 75th %iles) | N | Mean (SD) | Median (25th, 75th %iles) | Lower 95% CL for Mean | Upper 95% CL for Mean | Paired Test Used for Comparison | p-value |
| Pre-CPET PVP | mmHg | 26 | 9.8 (3.3) | 9.2 (7.4, 11.8) | 26 | 8.7 (2.9) | 7.9 (6.6, 10.9) | 26 | -1.1 (2.1) | -1.3 (-2.6, 0.4) | -2.001 | -0.269 | t-test | 0.012 |
| Post-CPET PVP | mmHg | 26 | 10.6 (3) | 9.3 (8.6, 13.2) | 26 | 9.5 (2.4) | 8.6 (8.1, 11) | 26 | -1.1 (2.1) | -1 (-1.8, 0.2) | -1.971 | -0.296 | t-test | 0.010 |
| Total Body Water (% of Total Weight) | % | 26 | 49.6 (8.3) | 50.1 (44.5, 56.4) | 26 | 48.4 (8.5) | 49 (42.6, 52.4) | 26 | -1.2 (4.7) | -0.5 (-1.1, 0.7) | -3.093 | 0.677 | signed rank test | 0.234 |
| Total Body Water (% of fat-free mass) | % | 26 | 72 (2.2) | 73.1 (70.1, 73.3) | 26 | 72.4 (2.4) | 73.2 (70.9, 73.4) | 26 | 0.4 (1.9) | 0 (-0.1, 0.1) | -0.353 | 1.184 | signed rank test | 0.290 |
| Measured Peak VO2 (ml/kg/min) | ml/kg/min | 26 | 19.5 (5.3) | 20.5 (16.2, 22.9) | 26 | 20.2 (5.1) | 21.5 (16, 22.8) | 26 | 0.7 (2) | 0.2 (-0.9, 2.2) | -0.048 | 1.532 | t-test | 0.064 |
| %Predicted VO2 Max | % | 26 | 52.5 (9.2) | 52.5 (48, 58) | 26 | 54.8 (8.3) | 52.5 (51, 63) | 26 | 2.3 (5.6) | 0.5 (-2, 7) | 0.093 | 4.599 | t-test | 0.042 |
| Measured O2 Pulse | ml/beat | 26 | 9.2 (2.2) | 8.9 (8.3, 9.7) | 26 | 9.6 (2.3) | 8.9 (8.1, 10.9) | 26 | 0.4 (1.1) | 0.3 (-0.6, 1.1) | -0.040 | 0.817 | t-test | 0.074 |
| %Predicted O2 Pulse | % | 26 | 73.7 (13.8) | 71.5 (65, 86) | 26 | 76.7 (12.9) | 78 (67, 82) | 26 | 3 (8.5) | 2 (-5, 11) | -0.425 | 6.425 | signed rank test | 0.133 |
| VE/VCO2 @AT |  | 25 | 33.1 (3.7) | 33 (30, 36) | 24 | 33.8 (4.1) | 33 (30, 37) | 24 | 1 (3) | 1 (-1, 3) | -0.308 | 2.225 | t-test | 0.131 |
| Respiratory Quotient Peak |  | 26 | 1.2 (0.1) | 1.2 (1.1, 1.3) | 26 | 1.2 (0.1) | 1.2 (1.1, 1.2) | 26 | 0 (0.1) | 0 (-0.1, 0) | -0.031 | 0.020 | t-test | 0.667 |
| OUES |  | 26 | 1342.5 (360.1) | 1372 (1120, 1506) | 26 | 1367 (426.6) | 1240 (1066, 1600) | 26 | 24.5 (204.5) | 16.5 (-130, 154) | -58.094 | 107.100 | t-test | 0.547 |
| ACHD PRO Summary Score |  | 26 | 86.2 (13.8) | 91.2 (82, 94.5) | 26 | 90.2 (10.3) | 92.8 (86.1, 97) | 26 | 4 (9.1) | 1.3 (-0.7, 6.5) | 0.268 | 7.648 | signed rank test | 0.016 |
| ACHD PRO Symptom Score |  | 26 | 84.5 (12.6) | 86.7 (74.7, 93.3) | 26 | 87.9 (12.3) | 90 (77.3, 100) | 26 | 3.4 (13.9) | 0 (-3.3, 6.7) | -2.185 | 9.005 | signed rank test | 0.447 |
| ACHD PRO Physical Limitation Score |  | 26 | 84.1 (17.4) | 90 (77.8, 100) | 26 | 88.6 (15) | 93.3 (86.7, 100) | 26 | 4.5 (6.4) | 1.7 (0, 6.7) | 1.858 | 7.052 | signed rank test | 0.001 |
| ACHD PRO Arrhythmia Score |  | 26 | 91.2 (13.3) | 97.2 (88.9, 100) | 26 | 91.9 (12.5) | 100 (86.7, 100) | 26 | 0.7 (13.2) | 0 (0, 5.6) | -4.593 | 6.046 | signed rank test | 0.879 |
| ACHD PRO Quality of Life Score |  | 26 | 87.7 (20.6) | 98.6 (81, 100) | 26 | 93 (12.9) | 100 (91.7, 100) | 26 | 5.2 (14.2) | 0 (0, 6) | -0.518 | 10.969 | signed rank test | 0.078 |
| ACHD PRO Anxiety/Depression Score |  | 26 | 83.5 (18.2) | 91.3 (75, 97.5) | 26 | 89.5 (13.5) | 95 (87.5, 100) | 26 | 6 (10.4) | 2.5 (0, 7.8) | 1.783 | 10.162 | signed rank test | 0.002 |

### Secondary Outcomes

Baseline post-exercise PVP decreased from 9.3mmHg (8.6, 13.2) to 8.6mmHg (8.1, 11; p=0.01; Table 2). Baseline peak VO2 was 20.5 ml oxygen/kg/min (16.2, 22.9) and had a trend toward an increase to 21.5 ml oxygen/kg/min (16, 22.8; p=0.06). Ve/VCO2, oxygen pulse, and oxygen uptake efficiency slope (OUES) did not significantly change during the study period. Total body water also did not change significantly during the study period. Baseline ACHD PRO V1 patient reported health status scores increased from 91.2 (82, 94.5) to 92.8 (86.1, 97; P=0.02). This change was predominantly due to improvement in the physical limitations (from 90 [77.8, 100] to 93.3 [86.7, 100] p<0.01) and depression/anxiety domains (from 91.3 [75, 97.5] to 95 [87.5, 100] p<0.01). When divided according to median baseline PVP, those with a PVP ≥ median had a trend toward increased peak VO2, decreased Ve/VCO2 and oxygen pulse. There was no difference between the groups in any of the other secondary outcomes (Figure 2).

### Safety and Laboratory Monitoring

Cystatin C (0.9mg/l [0.8, 0.9] - 0.9mg/l (0.8, 1); p<0.01), creatinine (0.8mg/dl [0.7, 0.9] - 0.9mg/dl [0.7, 1]; p=0.02) and blood urea nitrogen (14mg/dl [11, 17] – 15mg/dl [13, 18]; p=0.02) all increased during the study period while body weight (70.9kg [62.3, 83.7] – 70kg [62.7, 81.5]; p=0.02) and systolic blood pressure (112mmHg [108, 119] – 103mmHg [99, 114]; p=0.02) decreased (Table 3). There were no other changes in any safety parameters. Subjects generally tolerated the medications well with the most frequently reported side effects being increased urinary frequency or dizziness which did not affect participants’ usual daily activities (Table 4). There were no hospitalizations or deaths during the protocol. Pill counts can be found in Supplemental Table 2.

**Table 3:** Safety and Laboratory Monitoring; Significant values are indicated by italics.

| Safety and Laboratory Monitoring: Table 3 |  | Baseline |  |  | Follow-up |  |  | Change |  |  |  |  |  |  |
| --- | --- | --- | --- | --- | --- | --- | --- | --- | --- | --- | --- | --- | --- | --- |
|  | units | N | Mean (SD) | Median (25th, 75th %iles) | N | Mean (SD) | Median (25th, 75th %iles) | N | Mean (SD) | Median (25th, 75th %iles) | Lower 95% CL for Mean | Upper 95% CL for Mean | Paired Test Used for Comparison | p-value |
| Body Weight | kg | 26 | 72.2 (15) | 70.9 (62.3, 83.7) | 26 | 71.7 (14.6) | 70 (62.7, 81.5) | 26 | -0.5 (1.1) | -0.4 (-1.2, 0.3) | -0.9715971 | -0.0708312 | t-test | 0.025 |
| Height | cm | 26 | 168 (9.1) | 169.3 (160, 175.3) | 26 | 167.7 (8.6) | 169.6 (160, 174) | 26 | -0.3 (1.8) | 0 (-1.4, 0) | -1.034746 | 0.3832076 | signed rank test | 0.349 |
| Manual Systolic Blood Pressure | mmHg | 26 | 114 (13.7) | 112 (108, 119) | 26 | 107 (10.6) | 103 (99, 114) | 26 | -7 (13.9) | -7.5 (-18, 1) | -12.652044 | -1.4248789 | t-test | 0.016 |
| Manual Diastolic Blood Pressure | mmHg | 26 | 73.6 (10.5) | 71 (66, 82) | 26 | 69.3 (7.5) | 68 (66, 75) | 26 | -4.3 (11.1) | -2 (-8, 4) | -8.7605183 | 0.2220567 | t-test | 0.062 |
| Heart Rate per Minute |  | 26 | 82.4 (14.2) | 84.5 (71, 90) | 26 | 80.5 (16.7) | 83 (74, 90) | 26 | -1.9 (15.6) | -1.5 (-5, 7) | -8.2270799 | 4.380926 | signed rank test | 0.748 |
| Respiratory Rate per Minute |  | 26 | 14.9 (2.1) | 14 (14, 17) | 26 | 15.5 (2.5) | 15 (14, 18) | 26 | 0.6 (1.7) | 0 (0, 1) | -0.0708032 | 1.3015724 | signed rank test | 0.105 |
| Pulse Oximetry | % | 26 | 93 (3.3) | 93 (90, 95) | 26 | 92.1 (3.6) | 92 (90, 94) | 26 | -0.9 (3.8) | 0 (-4, 1) | -2.4512109 | 0.605057 | signed rank test | 0.368 |
| White blood count | K/cu mm | 25 | 6.3 (1.7) | 6.5 (5.2, 7.3) | 26 | 6.1 (1.7) | 6.5 (4.5, 7.1) | 25 | -0.3 (1.6) | -0.6 (-1.3, 0.1) | -0.9985274 | 0.3353274 | signed rank test | 0.087 |
| Red blood count | M/CU MM | 25 | 5.1 (0.5) | 5.1 (4.9, 5.4) | 26 | 5.2 (0.5) | 5.2 (4.9, 5.5) | 25 | 0.1 (0.3) | 0.1 (0, 0.3) | -0.0065436 | 0.2249436 | t-test | 0.063 |
| Platelet count | K/cu mm | 25 | 187.4 (53.7) | 160 (149, 225) | 26 | 184.1 (50.6) | 170.5 (149, 230) | 25 | -5.3 (25.6) | -3 (-14, 13) | -15.843935 | 5.2839353 | t-test | 0.313 |
| Hematocrit | % | 25 | 46.2 (4.4) | 46.6 (43.1, 49.2) | 26 | 47.1 (3.8) | 46.9 (43.7, 49.7) | 25 | 1 (2.8) | 1.4 (0, 2.5) | -0.2133944 | 2.1173944 | t-test | 0.105 |
| Sodium | mmol/L | 26 | 139.2 (1.8) | 139 (138, 140) | 26 | 139.1 (2.6) | 139.5 (138, 141) | 26 | -0.1 (2.4) | 0.5 (-2, 1) | -1.0591645 | 0.9053184 | signed rank test | 1.000 |
| Potassium | mmol/L | 25 | 4.1 (0.3) | 4.1 (4, 4.2) | 26 | 4.1 (0.4) | 4.1 (3.8, 4.3) | 25 | 0 (0.3) | 0 (-0.2, 0.2) | -0.087784 | 0.167784 | t-test | 0.524 |
| Chloride | mmol/L | 26 | 104.9 (3.6) | 104 (103, 108) | 26 | 104.6 (3.5) | 104.5 (102, 108) | 26 | -0.3 (2.4) | 0 (-1, 1) | -1.2955393 | 0.6801547 | t-test | 0.527 |
| CO2 | mmol/L | 26 | 23.8 (2.3) | 24 (23, 25) | 26 | 23 (2.8) | 23.5 (21, 25) | 26 | -0.7 (2.1) | -1 (-2, 1) | -1.5823 | 0.1207615 | t-test | 0.089 |
| Glucose | mg/dL | 26 | 87.5 (14.9) | 85 (81, 95) | 26 | 89.8 (15.1) | 89 (86, 96) | 26 | 2.3 (18.6) | 2.5 (-9, 16) | -5.222596 | 9.8379806 | t-test | 0.534 |
| BUN | mg/dL | 26 | 13.7 (3.9) | 14 (11, 17) | 26 | 15.4 (3.5) | 15 (13, 18) | 26 | 1.7 (3.5) | 1.5 (-1, 5) | 0.2318014 | 3.0758909 | signed rank test | 0.022 |
| Creatinine | mg/dL | 26 | 0.8 (0.2) | 0.8 (0.7, 0.9) | 26 | 0.9 (0.2) | 0.9 (0.7, 1) | 26 | 0 (0.1) | 0 (0, 0.1) | 0.0089376 | 0.0887547 | t-test | 0.018 |
| Calcium | mg/dL | 26 | 9.5 (0.3) | 9.5 (9.3, 9.8) | 26 | 9.6 (0.3) | 9.6 (9.4, 9.8) | 26 | 0.1 (0.3) | 0.1 (-0.1, 0.2) | -0.0428132 | 0.2043517 | t-test | 0.190 |
| Total Protein | g/dL | 26 | 7.4 (0.4) | 7.5 (7.1, 7.8) | 26 | 7.4 (0.5) | 7.5 (7, 7.9) | 26 | 0 (0.4) | 0.1 (-0.2, 0.3) | -0.1257356 | 0.1641971 | t-test | 0.787 |
| Albumin | g/dL | 26 | 4.5 (0.4) | 4.6 (4.2, 4.8) | 26 | 4.5 (0.4) | 4.5 (4.2, 4.8) | 26 | 0 (0.3) | 0.1 (-0.2, 0.2) | -0.1195125 | 0.0964356 | t-test | 0.828 |
| Bilirubin, Total | mg/dL | 26 | 0.8 (0.3) | 0.8 (0.6, 1) | 26 | 0.8 (0.3) | 0.8 (0.5, 1) | 26 | 0 (0.2) | 0 (-0.2, 0.1) | -0.0806691 | 0.1114384 | t-test | 0.744 |
| AST | U/L | 25 | 26.8 (7.8) | 26 (20, 32) | 25 | 28 (16.1) | 23 (21, 29) | 24 | 1.4 (14.1) | 0 (-5.5, 3) | -4.5249912 | 7.3583246 | signed rank test | 0.488 |
| ALT | U/L | 26 | 31.4 (15.6) | 27.5 (23, 35) | 26 | 32.8 (20) | 28.5 (23, 33) | 26 | 1.4 (12.6) | 1 (-5, 4) | -3.7092192 | 6.47845 | signed rank test | 0.923 |
| NT-Pro BNP | pg/mL | 14 | 175.8 (121.8) | 140 (106, 256) | 14 | 149.9 (90.8) | 127.5 (93, 210) | 14 | -25.9 (65.9) | -11 (-44, 11) | -63.986993 | 12.129851 | t-test | 0.165 |
| Cystatin C | mg/L | 26 | 0.9 (0.2) | 0.9 (0.8, 0.9) | 26 | 0.9 (0.2) | 0.9 (0.8, 1) | 26 | 0 (0.1) | 0 (0, 0.1) | 0.0224487 | 0.0767821 | signed rank test | 0.001 |

**Table 4:**
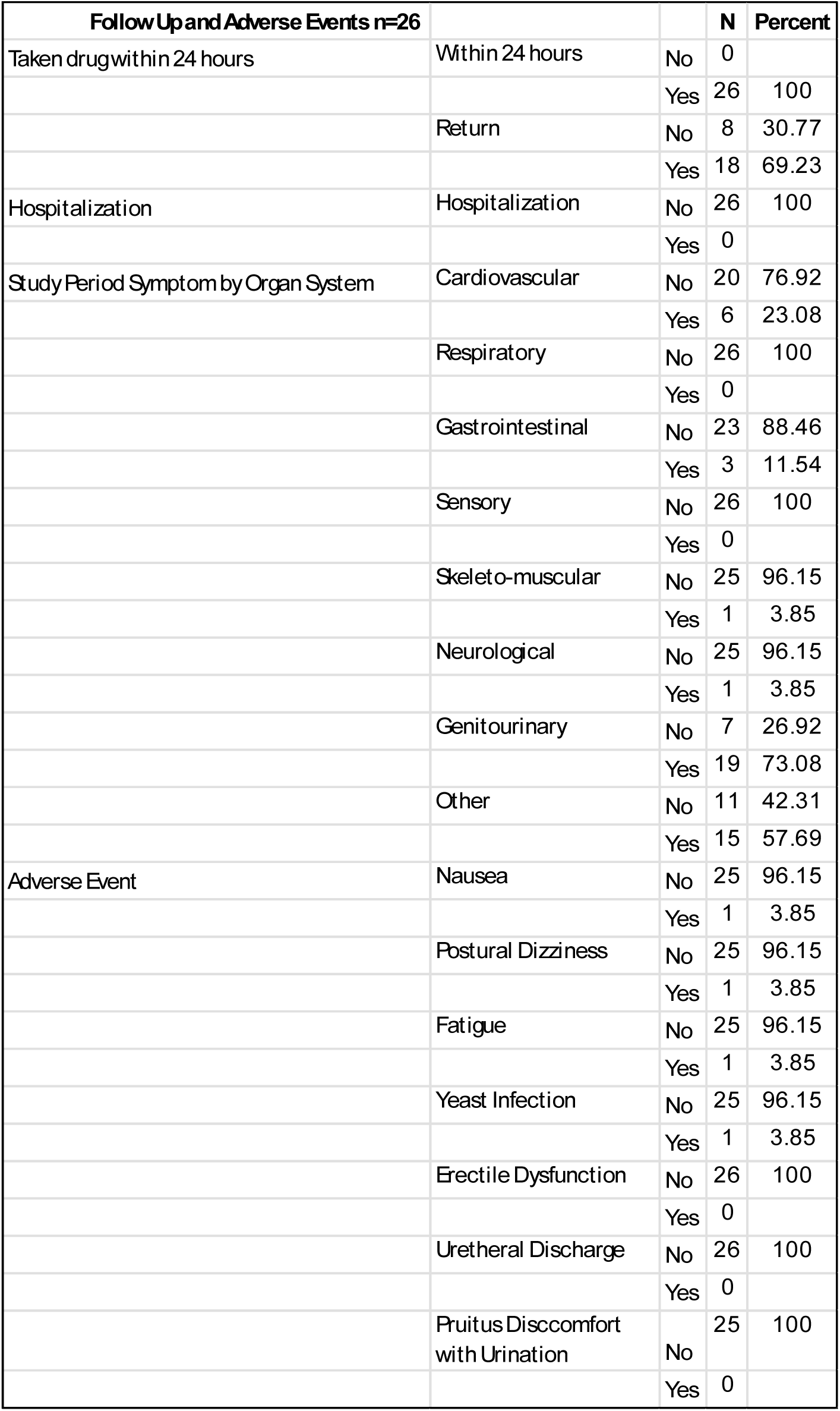
Follow up and Adverse Events; No caption.

| Table 4 - Follow Up and Adverse Events n=26 |  |  | N | Percent |
| --- | --- | --- | --- | --- |
| Taken drug within 24 hours | Within 24 hours | No | 0 |  |
|  |  | Yes | 26 | 100 |
|  | Return | No | 8 | 30.77 |
|  |  | Yes | 18 | 69.23 |
| Hospitalization | Hospitalization | No | 26 | 100 |
|  |  | Yes | 0 |  |
| Study Period Symptom by Organ System | Cardiovascular | No | 20 | 76.92 |
|  |  | Yes | 6 | 23.08 |
|  | Respiratory | No | 26 | 100 |
|  |  | Yes | 0 |  |
|  | Gastrointestinal | No | 23 | 88.46 |
|  |  | Yes | 3 | 11.54 |
|  | Sensory | No | 26 | 100 |
|  |  | Yes | 0 |  |
|  | Skeleto-muscular | No | 25 | 96.15 |
|  |  | Yes | 1 | 3.85 |
|  | Neurological | No | 25 | 96.15 |
|  |  | Yes | 1 | 3.85 |
|  | Genitourinary | No | 7 | 26.92 |
|  |  | Yes | 19 | 73.08 |
|  | Other | No | 11 | 42.31 |
|  |  | Yes | 15 | 57.69 |
| Adverse Event | Nausea | No | 25 | 96.15 |
|  |  | Yes | 1 | 3.85 |
|  | Postural Dizziness | No | 25 | 96.15 |
|  |  | Yes | 1 | 3.85 |
|  | Fatigue | No | 25 | 96.15 |
|  |  | Yes | 1 | 3.85 |
|  | Yeast Infection | No | 25 | 96.15 |
|  |  | Yes | 1 | 3.85 |
|  | Erectile Dysfunction | No | 26 | 100 |
|  |  | Yes | 0 |  |
|  | Urethral Discharge | No | 26 | 100 |
|  |  | Yes | 0 |  |
|  | Pruitus Discomfort with Urination | No | 25 | 100 |
|  |  | Yes | 0 |  |

## Discussion

In the present study, we investigated the effects of 4 weeks of daily dapagliflozin 10mg in a group of clinically stable adult patients with Fontan. We found that dapagliflozin decreased PVP by 1.3mmHg. While we found no significant impact on exercise parameters or total body water, we observed an improvement in patient-reported health status. The effects of dapagliflozin on the primary outcome were greatest in those with a higher baseline PVP. These data suggest that daily dapagliflozin may improve Fontan pressures and overall health status without compromising exercise capacity.

While the present study was of limited size, it suggests that the beneficial effects of dapagliflozin may be related to improvements in ventricular diastolic function. Although one of the proposed mechanisms underlying SGLT2i therapeutic benefit is a decrease in total body water^4^ we found no change in total body water in the present study. This may be due to the lack of change in diuretic dose in the present study, as loop diuretics appear to augment the impact of SGLT2 on this parameter ^9^. We also found a trend toward improved peak VO2 and oxygen pulse despite a decrease in PVP. These data indicate an increased stroke volume despite a decrease in filling pressures, which would be consistent with improved diastolic function. In view of these findings and the fact that the majority of the benefit observed was seen in those with preserved ventricular function, the present data suggest that dapagliflozin has a direct impact on myocardial relaxation in patients with Fontan. More granular physiologic data, however, are needed to substantiate this assumption.

Dapagliflozin appears to be generally well tolerated by patients with Fontan but some side effects bear monitoring. The majority of participants in the present study experienced no side effects and there were no hospitalizations or deaths. Patients in the study, however, experienced a decrease in body weight despite no change in total body water composition, suggesting somatic weight loss. Weight loss is a known side effect of dapagliflozin therapy^10^ likely due to calorie wasting. Given the propensity of patients with Fontan to sarcopenia^11^, broader utilization of dapagliflozin and SGLT2i generally in patients with Fontan would therefore merit monitoring for nutritional status.

In addition, we found an increase in both cystatin C and creatinine suggesting the possibility of a decrease in glomerular filtration rate (GFR). While the degree of change was not clinically significant and exercise parameters suggested an improvement in cardiac output with exercise, close monitoring of renal function after dapagliflozin initiation in patients with Fontan is likely warranted. The mechanism underlying this finding is unclear. Although this effect was not seen at study completion in the large seminal SGLT2i trials in heart failure^10,12^ an early reversible decrease in GFR upon SGLT2i initiation is known to occur in the heart failure population^13,14^ potentially due to (ultimately protective) decreases in glomerular perfusion pressures^15^. It is tempting to hypothesize that the decreases in systolic blood pressure and central venous pressure seen in the present study combined with afferent arteriolar vasoconstriction (related to increased sodium delivery to the macula densa), leading to a decreased GFR. If so, this would suggest dapagliflozin has similar renal protective effects in the Fontan population. Ultimately, longer-term studies will be needed to assess the trajectory of GFR changes in an SGLT2i-treated Fontan population over time.

The present data suggest that dapagliflozin could be a beneficial intervention to prevent long-term sequelae related to the Fontan circulation. While definitive longitudinal physiologic data are lacking, there is general belief that Fontan pressures rise over time due to progressive increases in pulmonary vascular resistance^16^ and ventricular diastolic dysfunction^17,18^. To date, pharmacologic interventions have focused largely on decreasing pulmonary vascular resistance^19–21^. In the present study, we aimed to improve diastolic function. If the decreases in Fontan pressure demonstrated in the present study can be sustained over time, there is the potential that targeted therapy with dapagliflozin and pulmonary vasodilators^22,23^ may represent the beginnings of a Fontan-specific therapeutic cocktail, analogous to that which has been beneficial in heart failure with normal cardiac anatomy. While these drugs do not reverse the unfavorable Fontan physiology, early and sustained utilization may delay deterioration and targeted use in current patients may turn back the clock on the Fontan circulation, prolonging the window preceding Fontan failure.

The present study has multiple limitations. The study size was small and study design was unblinded creating the possibility that either unmeasured variables or study population selection were responsible for the present findings. In particular, the unblinded design may have had an impact on patient reported health status. Any improvement in exercise capacity over the course of the study could also be attributable to training effect with repeated testing. As a pilot study, all present findings therefore bear confirmation in a larger blinded study.

In conclusion, the present data suggest that dapagliflozin may be beneficial in reducing Fontan pressures and improving health status without compromising exercise capacity in adult patients with Fontan and elevated PVP.

## Data Availability

Data will be made available upon written request to the corresponding author

---

**Supplemental Table 1:** Concomitant medications.

| **Supplemental Table 1:**<br>**Concomitant medications** |  |  |  |  |
| --- | --- | --- | --- | --- |
|  |  |  | **N** | **Percent** |
|  | Diuretics | No | 17 | 65.38 |
|  | PDE5 Inhibitors | No | 24 | 92.31 |
|  | Aspirin | No | 7 | 26.92 |
|  | Anticoagulants | No | 18 | 69.23 |
|  | Antiarrhythmics | No | 24 | 92.31 |
|  | Calcium Channel Blockers | No | 26 | 100 |
|  | Beta-blockers | No | 21 | 80.77 |
|  | ACE Inhibitors or ARBs | No | 12 | 46.15 |
|  | Mineralo-corticoid Receptor Antagonists | No | 17 | 65.38 |
|  | Digoxin | No | 24 | 92.31 |
|  | Other | No | 10 | 38.46 |

**Supplemental Table 2.** Pill count.

| **Supplemental Table 2** |  | **Leftover pills** | **N** | **Percent** |
| --- | --- | --- | --- | --- |
| Pill count | Leftover pill | 0 | 1 | 3.85 |
|  |  | 1 | 3 | 11.54 |
|  |  | 2 | 12 | 46.15 |
|  |  | 3 | 6 | 23.08 |
|  |  | 4 | 2 | 7.69 |
|  |  | 5 | 1 | 3.85 |
|  |  | 8 | 1 | 3.85 |

